# Transcriptomics-Guided Drug Repurposing Identifies Candidate Compounds for Improving Long-Term Stroke Outcome

**DOI:** 10.64898/2026.03.09.26347942

**Authors:** Natalia Cullell, Cristina Gallego-Fábrega, Jara Cárcel, Elena Muiño, Laia Llucià-Carol, Jesús M Martín-Campos, Israel Fernández-Cadenas, Jerzy Krupinski

**Affiliations:** Stroke Pharmacogenomics and Genetics group, Biomedical Research Institute Sant Pau (IIB Sant Pau). 08041. Barcelona; Neurology. Hospital Universitari MútuaTerrassa/ Fundacio Docència i Recerca MutuaTerrassa, 08221. Terrassa, Spain; Facultat de Medicina. Universitat de Barcelona, 08036. Barcelona, Spain

## Abstract

**Background:** Functional recovery after ischemic stroke is a complex and polygenic process influenced by genetically regulated molecular pathways. Although genome-wide association studies (GWAS) have identified variants associated with stroke outcome, translating these findings into actionable therapeutic targets remains challenging. Transcriptome-wide association studies (TWAS) provide a framework to link genetic variation to gene expression and functional phenotypes, enabling mechanism-driven drug repurposing strategies.

**Objective:** To identify candidate compounds capable of improving long-term functional outcome after ischemic stroke by integrating GWAS summary statistics with brain transcriptomic data and large-scale perturbational drug signatures.

**Methods:** We performed TWAS integrating summary statistics from a stringent GWAS of long-term stroke outcome (modified Rankin Scale at three months, mRS3; N = 1,791; 8,895,027 variants) from the GODS study with brain eQTL data from ten regions in the GTEx project. Pathway enrichment analysis was conducted using WebGestalt. Drug repurposing was performed using the Trans-phar pipeline by comparing TWAS-derived transcriptional signatures with compound-induced gene expression profiles from the Connectivity Map (CMap) L1000 dataset across five neural cell lines. Compounds showing inverse transcriptional correlations were prioritized and further evaluated based on existing clinical and preclinical evidence in stroke.

**Results:** TWAS identified 22 genes consistently ranked within the top 10% across all ten regions. Pathway enrichment analysis highlighted transcriptional regulation processes, with the RNA polymerase pathway reaching statistical significance after FDR correction in the broader gene set. Drug repurposing analysis identified nine compounds whose transcriptional signatures inversely correlated with genetically predicted expression profiles associated with poor functional outcome. Among these, anandamide and progesterone had prior clinical evidence in stroke, while Z-guggulsterone demonstrated preclinical neuroprotective potential.

**Conclusions:** Integration of GWAS and brain eQTL data identifies robust transcriptional signatures associated with long-term stroke outcome and supports transcriptomics-driven drug repurposing strategies. This human genetics–guided framework prioritizes candidate compounds with potential translational relevance for improving functional recovery after ischemic stroke.

## INTRODUCTION

Stroke remains one of the leading causes of long-term disability worldwide (1). Although considerable progress has been made in understanding the genetic determinants of stroke risk (2), substantially less is known about the biological mechanisms influencing long-term functional outcome after ischemic stroke. Functional recovery is a highly complex and polygenic process involving neuronal survival, neuroinflammation, synaptic remodeling, and repair pathways (REF), many of which are under genetic regulation.

Genome-wide association studies (GWAS) have identified genetic variants associated with functional outcome, both at initial (NIHSS24h) (3) and long-term (using modified rankin scale at three months: mRS3) (3,4); however, translating these associations into actionable therapeutic targets remains challenging. Transcriptome-wide association studies (TWAS) enable the integration of GWAS summary statistics with expression quantitative trait loci (eQTL) data, thereby identifying genetically regulated gene expression associated with disease phenotypes (5). Few studies have applied this approach in stroke, and they have focused only on stroke risk. The cis-regulation at the transcriptomic level of 19 genes in blood and adipose tissue were identified by Yang et al (6) in association with stroke risk. Traylor et al. (7), found six genes using TWAS whose expression was regulating small vessel stroke (SVS) risk. This approach provides mechanistic insight into how inherited variation may influence post-stroke recovery.

Despite decades of research, neuroprotection in ischemic stroke remains an unresolved challenge. More than one thousand compounds have demonstrated neuroprotective effects in preclinical animal models, yet none has shown consistent clinical benefit in phase III trials (8). Several factors likely contribute to these translational failures, including biological differences between animal models and human stroke pathology, patient heterogeneity, narrow therapeutic time windows, and the lack of biomarker-based stratification strategies (9). These limitations highlight the need for alternative, mechanism-driven approaches that leverage human genetic data to prioritize therapeutic targets with higher translational potential.

Recent advances in human genetics have highlighted the value of genetically informed approaches to identify therapeutic targets for complex diseases. An illustrative example in the field of secondary prevention of ischemic stroke is the identification of coagulation factor XI inhibition as a potential therapeutic strategy. Integrative genomic approaches, including Mendelian randomization analyses combining genetic and proteomic data, have provided causal evidence supporting the role of factor XI in thrombotic risk (10). These findings contributed to the development of factor XI inhibitors such as asundexian, which has recently been evaluated in the OCEANIC clinical trial in patients with recent ischemic stroke or high-risk TIA (11). The successful translation of genetic discoveries into clinical trials underscores the potential of integrative multi-omic approaches to identify biologically supported therapeutic targets and facilitate the development of novel strategies for stroke.

Drug repurposing strategies based on transcriptomic signatures offer an efficient framework to identify compounds capable of modulating disease-associated gene expression profiles. By comparing disease-related transcriptional signatures with large-scale perturbational datasets, it is possible to prioritize compounds whose effects oppose pathological gene expression patterns (12). In this study, we applied Trans-phar (13), a transcriptomics-driven drug repurposing pipeline, to identify bioactive compounds in long-term ischemic stroke outcome. By integrating GWAS summary statistics for long-term stroke outcome with brain eQTL data across multiple regions and comparing the resulting TWAS-derived signatures with transcriptional profiles from brain-derived cell lines treated with thousands of bioactive compounds, we aimed to identify compounds capable of reversing genetically regulated expression patterns associated with worse functional recovery.

## METHODS

### Study design

This was a transcriptomics-based drug repurposing study integrating long-term (mRS3) stroke outcome GWAS, brain eQTL data, and compound-induced transcriptional signatures through the Trans-phar pipeline (13).

### Genomic data

Summary statistics from the long-term stroke outcome GWAS evaluating functional status at three months (mRS3) were used. Among the analyses available from the Genetics of Ischaemic Stroke Functional Outcome (GODS) study, the stringent meta-analysis was selected (14). This analysis included 1,791 subjects from seven independent cohorts comprising patients with anterior circulation ischemic stroke who were functionally independent before stroke (mRS <3). The GWAS model was adjusted for NIHSS at discharge, stroke etiology, sex, smoking habit, and two principal components. The summary statistics included 8,895,027 single nucleotide variants.

### eQTL data

Brain eQTL summary statistics were obtained from the Genotype-Tissue Expression (GTEx) Project (version 7) (15). Ten brain regions were analyzed: anterior cingulate cortex, caudate basal ganglia, cerebellar hemisphere, cerebellum, cortex, frontal cortex, hippocampus, hypothalamus, nucleus accumbens, and putamen.

Transcriptome-wide association studies were performed separately for each brain region by integrating GWAS summary statistics with region-specific eQTL data. For each region, genes were ranked according to the absolute value of the TWAS Z-score, and the top 10% were selected for downstream analyses, representing genes with the strongest genetically regulated differential expression associated with long-term stroke outcome.

To prioritize genes with consistent cross-regional signals, we evaluated the overlap of top-ranked genes across tissues. Genes present within the top 10% in more than five of the ten brain regions were considered broadly represented, while genes consistently ranked within the top 10% across all regions were considered highly robust candidates.

### Pathway enrichment analysis

Pathway enrichment analyses were conducted using KEGG pathway database (16) in WebGestalt (17). False discovery rate (FDR) correction was applied to account for multiple testing.

### Drug repurposing analysis

Drug repurposing analyses were performed using transcriptional signatures from the Connectivity Map (CMap) L1000 dataset (18) with Trans-Phar. Compound-induced gene expression profiles were obtained from motor neuron–enriched cells (MNEU.E), neuronal cells (NEU), neural progenitor cells (NPC), neural progenitor cells with Cas9 (NPC.Cas9), and neural progenitor cells with TAK (NPC.Tak). The number of unique compounds per cell line ranged from 61 to 2,346 depending on the cell type, and when considering different doses and timepoints separately, a total of 165,361 tissue–cell line–compound combinations were evaluated.

Within the Trans-phar framework, the top 10% TWAS-ranked genes from each brain region were correlated with compound-induced gene expression signatures using negative Spearman rank correlations. Compounds showing inverse transcriptional profiles relative to the genetically predicted expression associated with worse stroke outcome were prioritized as potential therapeutic candidates.

Candidate drugs were further prioritized based on existing evidence from preclinical or human clinical trials in stroke registered in ClinicalTrials.gov (19).

## RESULTS

Integration of long-term stroke outcome GWAS data with brain eQTL datasets through TWAS identified a substantial number of genes with genetically regulated expression associated with functional recovery (mRS3). The number of genes within the top 10% ranked by absolute Z-score varied across brain regions, ranging from 361 genes in the hippocampus to 807 genes in the cerebellum. Specifically, 403 genes were identified in the anterior cingulate cortex, 543 in the caudate basal ganglia, 807 in the cerebellum, 643 in the cerebellar hemisphere, 467 in the frontal cortex, 361 in the hippocampus, 367 in the hypothalamus, 487 in the nucleus accumbens, 593 in the cortex, and 428 in the putamen.

Cross-regional comparison revealed that 140 genes were present within the top 10% in more than five brain regions, representing over half of the tissues analyzed. Notably, 22 genes were consistently ranked within the top 10% across all ten brain regions, indicating highly robust and region-independent associations with long-term stroke outcome (Table 1).

**Table 1:**
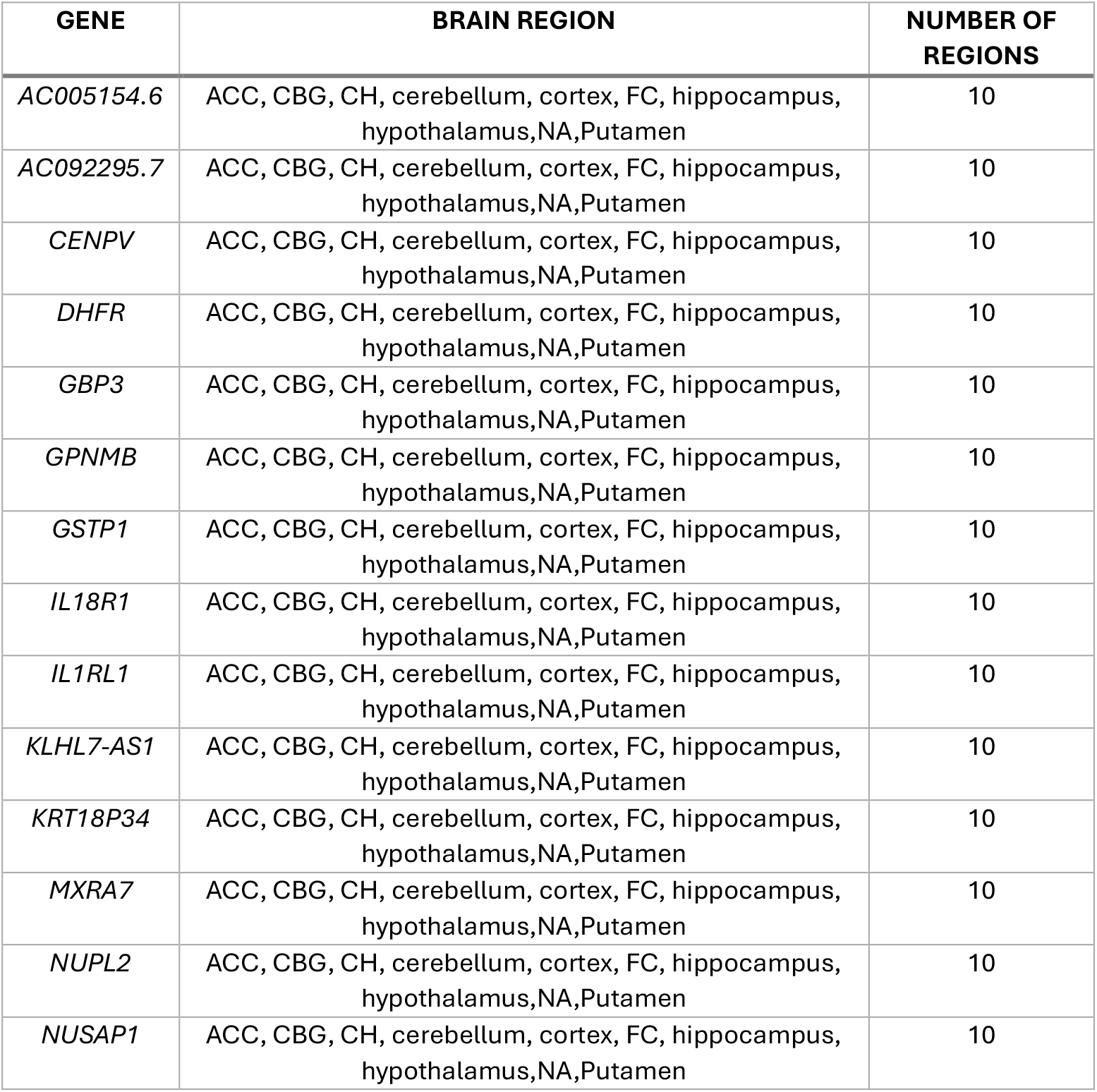
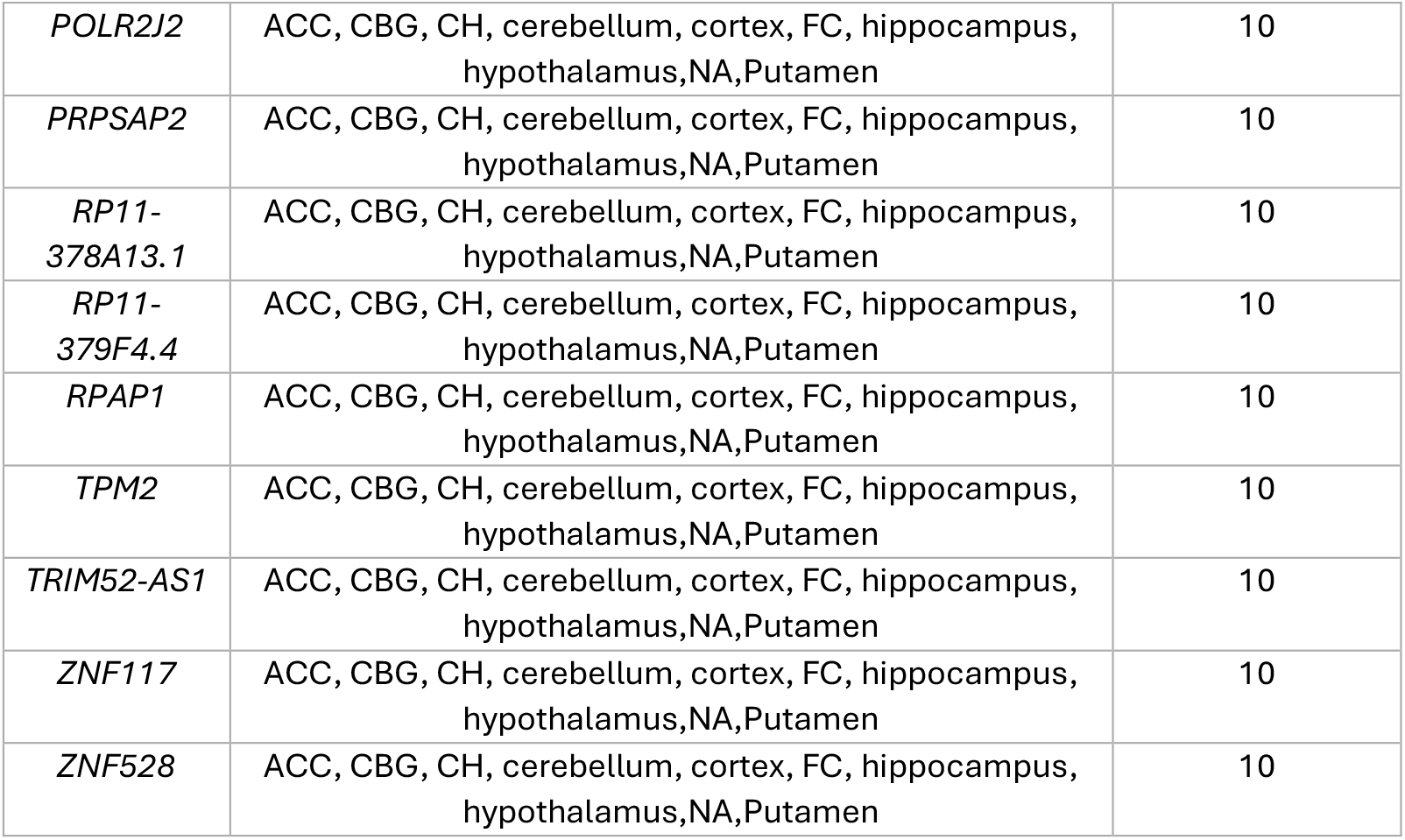
Top 10% genes with differential gene expression associated with long-term outcome in all the studied brain regions. ACC: Anterior cingulate cortex; CBG: caudate basal ganglia; CH: cerebellar hemisphere; FC: frontal cortex, NA: nucleus accumbens

Pathway enrichment analysis of the 22 consistently ranked genes did not reveal pathways reaching statistical significance after false discovery rate (FDR) correction; however, several biologically relevant pathways showed nominal associations (p < 0.05), including folate biosynthesis, RNA polymerase, drug metabolism, and immune-related pathways (Figure 1A). When expanding the analysis to the 140 genes present in more than 50% of brain regions, similar biological themes were observed. Importantly, in this broader gene set, the RNA polymerase pathway reached statistical significance after FDR correction (Figure 1B), suggesting that transcriptional regulation mechanisms may play a relevant role in long-term functional recovery after ischemic stroke.

**Figure 1.**
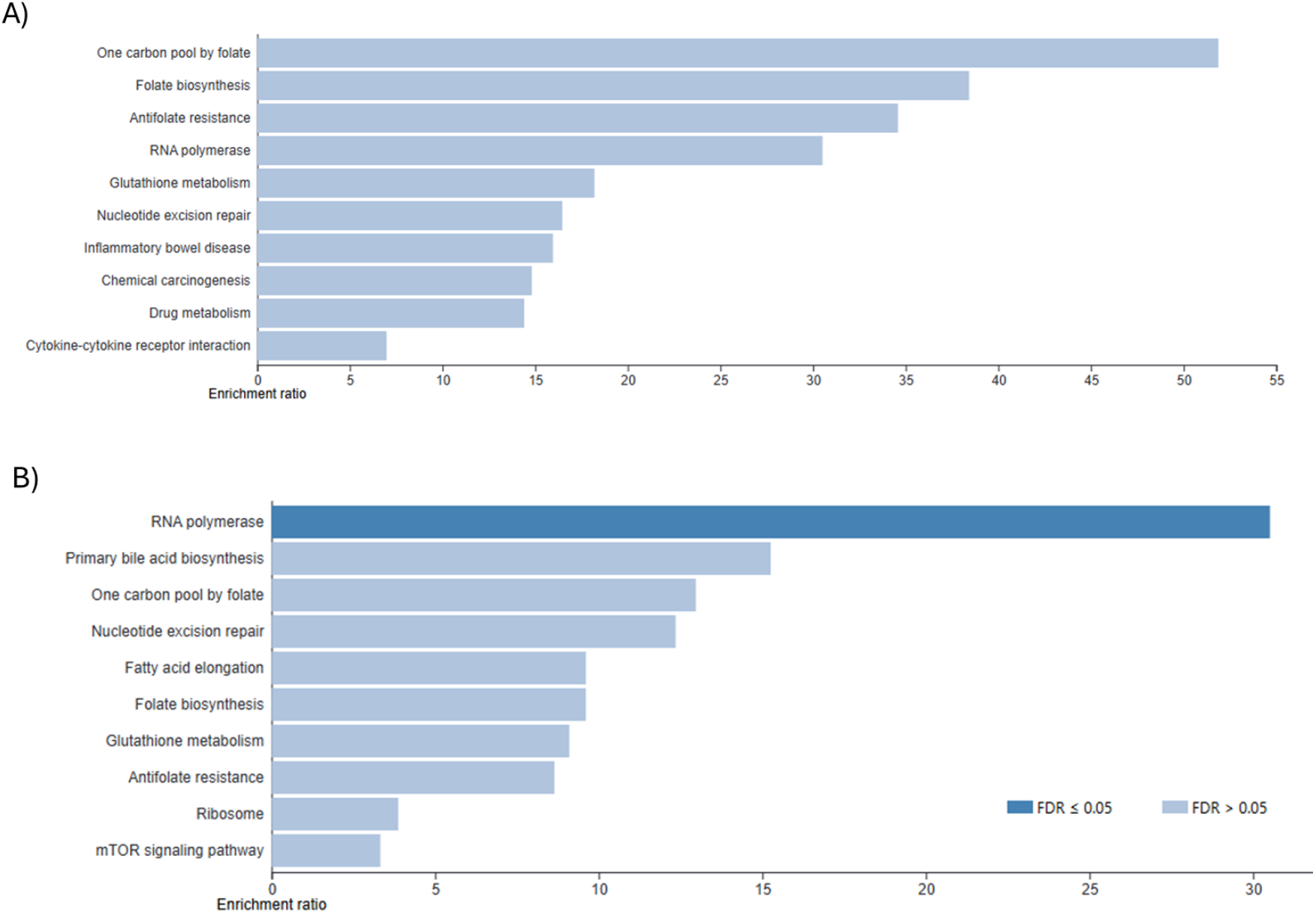
Pathway enrichment analysis A) analysis for the 140 genes belonging to the top 10% differentially expressed genes in all the studied brain regions and B) for the 140 genes belonging to the top 10% differentially expressed genes in more than 50% of the studied brain regions.

Drug repurposing analysis using Trans-phar systematically evaluated 165,361 combinations of brain region, cell line, and compound condition. By identifying compounds whose transcriptional signatures inversely correlated with the genetically predicted gene expression associated with worse stroke outcome, we prioritized candidate bioactive compounds with potential neuroprotective properties for long-term functional recovery after ischemic stroke.

A total of nine compounds were identified whose transcriptional regulation in neuronal cell lines showed an inverse pattern compared with the genetically predicted expression profile associated with poor stroke outcome (Table 2). These candidates were further filtered according to available clinical and preclinical evidence in stroke. Among them, anandamide and progesterone had some degree of clinical evidence, although limited. Z-guggulsterone showed preclinical evidence of neuroprotective effects but lacked clinical validation in stroke patients.

**Table 2:** Drug repurposing results with Trans-phar. Motor neuron–enriched cells (MNEU.E), neuronal cells (NEU), neural progenitor cells (NPC), neural progenitor cells with Cas9 (NPC.Cas9), and neural progenitor cells with TAK (NPC.Tak)

| Brain_Region | Drug | Cell_line | Rho | P-value |
| --- | --- | --- | --- | --- |
| Hypothalamus | anandamide | NEU | -0,3110 | 5,57E-04 |
| Anterior_cingulate_cortex | etoposide | NPC.TAK | -0,3218 | 1,06E-04 |
| Anterior_cingulate_cortex | huperzine | NEU | -0,3112 | 1,75E-04 |
| Hypothalamus | osimertinib | NPC | -0,3059 | 6,77E-04 |
| Anterior_cingulate_cortex | pazopanib | NEU | -0,3067 | 2,15E-04 |
| Anterior_cingulate_cortex | progesterone | NPC | -0,3180 | 1,27E-04 |
| Anterior_cingulate_cortex | rebastinib | NPC | -0,3203 | 1,14E-04 |
| Hypothalamus | rotenonic.acid | NEU | -0,3063 | 6,68E-04 |
| Nucleus_accumbens_basal_ganglia | z.guggulsterone | NPC | -0,3159 | 1,99E-05 |

## DISCUSSION

In this transcriptomics-driven drug repurposing study integrating long-term stroke outcome GWAS data with brain eQTL datasets and large-scale perturbational signatures, 22 genes were consistently ranked within the top 10% across all analyzed brain regions, and 140 genes were shared across more than half of the tissues. Pathway enrichment analyses highlighted transcriptional regulation mechanisms, particularly the RNA polymerase pathway, as significantly enriched after FDR correction in the broader gene set. These findings suggest that genetically regulated transcriptional machinery and downstream regulatory networks may play a central role in post-stroke recovery.

Importantly, the drug repurposing strategy prioritized a total of nine candidate compounds with potential neuroprotective properties in ischemic stroke whose transcriptional signatures inversely correlated with these genetically predicted patterns associated with poor outcomes, thereby providing mechanistic coherence between genetic architecture and pharmacological modulation. Among them, progesterone, anandamide, huperzine A, and Z-guggulsterone emerged as particularly relevant candidates due to previous biological evidence and/or clinical evaluation in neurological conditions.

Among the prioritized compounds, progesterone stands out as a biologically plausible neuroprotective candidate. Progesterone is a steroid hormone synthesized both in peripheral tissues, such as the ovaries and adrenal glands, and locally in the brain as a neurosteroid (20). Preclinical studies have consistently demonstrated neuroprotective effects of progesterone in ischemic stroke models, including reductions in infarct volume (ranging from 30% to 50% in animal models), attenuation of cerebral edema, decreased neuroinflammation and neuronal apoptosis, improved functional recovery, and an extended therapeutic window of up to 24 hours post-ischemia (21–23). Mechanistically, progesterone modulates inflammatory cascades, reduces oxidative stress, stabilizes mitochondrial membranes, regulates apoptotic pathways, and promotes remyelination and repair processes (24).

Clinical translation of progesterone has primarily been explored in traumatic brain injury (25) through large trials such as ProTECT (26) and SyNAPSe (27). Although early-phase studies suggested safety and potential benefit, phase III trials yielded inconsistent results, likely due to patient heterogeneity, lack of biomarker-based stratification, and timing variability. Importantly, no previous clinical study has evaluated progesterone in ischemic stroke with genetic or transcriptomic stratification. Our results provide a rationale for revisiting progesterone within a precision medicine framework guided by genetically regulated expression signatures.

Anandamide, an endogenous endocannabinoid (28), was also identified as a candidate compound. The endocannabinoid system is increasingly recognized as a regulator of neuroinflammation, excitotoxicity, and synaptic plasticity, all central mechanisms in stroke pathophysiology (29). Anandamide levels have been associated with stroke prognosis in observational studies, suggesting a potential endogenous compensatory response (30). A clinical trial using Sativex, a cannabinoid extract, has been conducted in stroke patients, although focused on spasticity rather than neuroprotection (31). While direct interventional evidence for anandamide in acute ischemic stroke remains limited, the convergence of observational prognostic associations and transcriptomic inverse correlation in our study supports further mechanistic exploration.

Huperzine A, a naturally occurring alkaloid marketed as a dietary supplement, has been investigated primarily in Alzheimer’s disease for cognitive enhancement (32), with mixed or negative results in large trials. A clinical study in hypertensive intracerebral hemorrhage has also been reported (33). Huperzine A exerts acetylcholinesterase inhibition and may modulate neuroinflammatory and oxidative pathways (34), which could intersect with post-stroke recovery mechanisms. However, its over-the-counter availability and heterogeneous regulatory status warrant careful evaluation before clinical translation.

Z-guggulsterone, a bioactive component derived from traditional medicinal preparations used in China and India (35), has shown preclinical evidence of neuroprotective effects (36). Recent single-cell transcriptomic studies have suggested regulation of Spp1 (osteopontin), a gene implicated in neuroinflammation and repair processes (37). Despite the absence of clinical trials in stroke, its molecular plausibility and transcriptomic inverse signature in our analysis make it an intriguing exploratory candidate.

Collectively, our findings highlight two major conceptual advances. First, they reinforce the importance of transcriptional regulation pathways, particularly RNA polymerase-related mechanisms, in long-term stroke recovery. Second, they demonstrate the feasibility of integrating human genetic data with perturbational transcriptomics to prioritize compounds with mechanistic alignment to genetically driven disease biology. This approach addresses one of the key limitations of previous neuroprotection strategies: the lack of human genetic anchoring in therapeutic selection.

Nevertheless, several limitations must be acknowledged. TWAS relies on genetically predicted expression and does not capture dynamic post-stroke transcriptional changes driven by environmental or temporal factors. The use of GTEx brain tissues, derived from non-stroke donors, may not fully reflect post-ischemic transcriptional states. Additionally, compound signatures were obtained from in vitro cell lines, which cannot recapitulate the full complexity of the ischemic brain microenvironment, including vascular, immune, and systemic interactions.

In conclusion, this integrative multi-omic approach identified nine candidate compounds with potential neuroprotective effects in ischemic stroke, including progesterone and anandamide, which have prior clinical or biological support. Our findings provide a genetically informed framework for re-evaluating neuroprotective therapies and support the development of precision medicine strategies in stroke recovery.

## Acknowledgments

We thank the International Stroke Genetics Consortium, the Spanish Stroke Genetics Consortium, the International Stroke Genetics Consortium, and the Global Alliance for Stroke acute and long-term outcome genetics and the RICORS Network (RD24/0009/0029).

## Author Contributions

I.FC. and J.K. are responsible for the study design and the analytical accuracy of the manuscript. N.C. performed statistical analyses and prepared the manuscript. C.GF., J.CM., L.LLC, E.M. and J.K reviewed the manuscript and contributed to the data interpretation. All the authors reviewed and approved the manuscript content.

## Institutional Review Board

The study was conducted according to the guidelines of the Declaration of Helsinki. The different studies included in this manuscript were responsible for the aprovement by the Institutional Review Board.

## Informed Consent

All the studies included as part of this article were responsible for the obtention of the informed consent of the patients.

## Data Availability

The datasets used and/or analysed during the current study are available from the corresponding author on reasonable request.

